# Effectiveness of Psychosocial care interventions in improving the Quality of Life for Adult Patients with Cancer in low-and middle-income countries (LMICs): A Systematic Review and Meta-Analysis Protocol

**DOI:** 10.1101/2024.02.08.24302499

**Authors:** David Kavuma, Eve Namisango, Julia Downing, Nixon Niyonzima, Alison Annet Kinengyere, Simon Kasasa, Fredrick Edward Makumbi, Ekwaro Anthony Obuku, Fred Nuwaha

## Abstract

**Background:** Psychosocial care interventions are part and parcel of cancer care and are known for their significant contribution to the improvement of the quality of life (QoL) for cancer patients and their families. Assessment of the QoL of patients with cancer and their families has become critical in cancer care nowadays since it guides health care providers in making informed decisions during the care process. The aim of this meta-analysis is to synthesise the online literature of primary studies from LMICs in order to understand the effectiveness of psychosocial care interventions towards the improvement of the QoL of adult patients with cancer.

**Methods:** This study will be done in tandem with the Preferred Reporting Items for Systematic Reviews and Meta-Analyses (PRISMA) standards. Primary studies that would have investigated the effectiveness of psychosocial care interventions on quality of life of adult cancer patients will be identified through searches in the various electronic databases that are known to produce optimal and efficient searches for systematic reviews and meta-analyses include: Ovid MEDLINE(R), PubMed, EMBASE, APA PsychINFO, Web of Science, PubMed, and Google Scholar. Studies published between 1^st^ January 2002 and 31^st^ December 2023 in any LMIC, will be searched. After developing the meta-analysis question, we have developed a search string from the PICOST (population, intervention, comparison, outcome, study design, setting and Time-frame) model with a limitation on study design.

**Discussion:** This systematic review and meta-analysis will gather evidence from primary studies on the effectiveness of psychosocial care interventions in improving the QoL for adult patients with cancer in LMICs.

**Protocol registration:** This protocol was registered on 5^th^ June 2023 and its registration number is CRD42023421561.

## 1.0 Introduction

The Global cancer (GLOBOCAN) statistics indicate that one in 8 men and one in 11 women died from cancer in 2018(1) and that the International Agency for Research on Cancer (IARC), notes that, one in every six deaths is due to cancer with 70% of deaths from cancer occur LMICs(2). WHO projects that the incidence of cancer will exponentially increase by the year 2030, with an annual number of cancer cases rising from 14.1 million in 2012 to 21.6 million in 2030 and deaths due to cancer rising from 8.8 million worldwide in 2015 to over 12 million in 2030(3).

Available evidence indicates that cancer is the most feared disease(4). This is the reason as to why being diagnosed with cancer, is indeed a very frightening experience with significant impact on the patient, caregivers, friends, family members and the community from diagnosis till end of life(5). Apart from physical suffering during diagnosis, surgery, chemotherapy or radiotherapy, cancer also affects the patient’s psychological dimension (characterized by shock, mental distress, anxiety, hopelessness, anger, depression, reduced self-esteem, fear of possible death to happened soon), the social dimension (interruption in participation in social or community activities, depending on others, loneliness and abandonment) and spiritual dimension (like loss of hope and faith, change in one’s perspective of life and existence)(3,6). This is the reason as to why cancer care should not only utilise biomedical interventions (medical, radiotherapy and surgery) to address the health needs of cancer patients since their needs are diverse and require holistic approach(6). Health care providers should always integrate psychosocial care with biomedical interventions in routine cancer care for adult patients with cancer in order to holistically address their comprehensive needs patients for optimal improvement of their QoL(6–10). This is because, even as recognized by WHO that, psychosocial well-being is key component of complete health(11) thus, it should be promoted throughout the continuum of cancer care.

Relatedly, the Worldwide Hospice Palliative Care Alliance (WHPCA) and WHO emphasise that, all interventions intended to reduce suffering among people with life-threatening illnesses, like cancer, should integrate psychosocial care so as to provide relief from pain and distressing symptoms, thereby improving the QoL of patients with cancer and their families(6). In 2014, after recognizing the persistent global rise in non-communicable and other chronic diseases, the Sixty-seventh World Health Assembly (WHA) approved a resolution to strengthen care of patients with non-communicable diseases, and governments were urged to strengthen integration of such care in their health care systems at all levels(12).

Following this declaration, most countries have continued to strengthen the integration of cancer care(13). By standards, cancer care is incomplete without psychosocial care(14). So, with the 2014 WHA resolution, it is implied that psychosocial care is integrated in cancer care. Correspondingly, available literature indicate that a lot of original research on psychosocial support in cancer care has been done in developed countries and quite limited ones in developing countries. To the best of our knowledge, there is scarcity of evidence on systematic reviews and meta-analyses on psychosocial care among adult patients with cancer in LMICs. Secondly, no systematic review and meta-analysis has been done on the effectiveness of psychosocial care interventions among adult patients with cancer in LMICs. This systematic review and meta-analysis will collate, summarise and sythesise what is already known from original research on the effectiveness of psychosocial care interventions among adult patients with cancer care in LMICs. The results of this study will contribute to the development of the future agenda for psychosocial support in cancer care in LMICs where the cancer burden continues to rise each year.

### 1.1 Systematic Review question

This systematic review and meta-analysis will be guided by this question, ‘what research exists online relating to the effectiveness of psychosocial interventions in the improvement of the quality of life of adult patients with cancer in LMICs?’ This question will be used as a basis for generating the search terms. Thus, the research question will give valuable and relevant evidence about the systematic review and meta-analysis while addressing the specific elements of PICOST, where P=Population (adult patients with cancer), I=Intervention (psychosocial care interventions), C=Comparison (Comparison of the study group receiving the intervention with the one receiving the usual cancer care), O=Outcome (Measurement of the improvement (effect) caused by the intervention on the quality of life (QoL) in intervention group), Study design (Randomised controlled trial, quasi-experimental studies, cohort studies controlled before-and-after studies and other designs with comparison group, S=Setting (Low-and middle-income countries) and T=Time frame (January 2002 to December 2023).

### 1.2 Context (Setting)

This systematic review and meta-analysis will include original studies carried out in Low-and middle-income countries (LMICs) as listed by World Bank (Appendix 1).

### 1.3 Rationale

The primary objective of this systematic review and meta-analysis is to investigate the effectiveness of psychosocial care interventions in the improvement of the quality of life of adult patients with cancer versus the standard or usual cancer care in LMICs.

### 1.4 Objective

This systematic reviews and meta-analysis intends to determine the effectiveness of psychosocial care intervention in the improvement of the quality of life of adult patients with cancer compared with usual cancer care in LMICs.

### 1.5 Definition of Health related Quality of Life

WHO defines health-related quality of life (HRQOL) as the wide range of human experiences that include physical, role, emotional, cognitive, and social functioning(15) which were anchored on the WHO definition of health(16). The domains of this systematic review and meta-analysis will be attached to elements of HRQOL as defined by WHO.

### 1.6. Definition of Psychosocial care

The National Council for Hospice and Specialist Palliative Care Services defines psychosocial care as a practice that is concerned with psychological, social and emotional well-being of the patient and their family(17). In addition, psychosocial care incorporates spiritual or pastoral care services, adaptation to the illness and its effects as well as therapeutic communication issues(18).

## 2.0 Methods

### 2.1 Information sources

This study will be done in tandem with the Preferred Reporting Items for Systematic Reviews and Meta-Analyses Protocol (PRISMA-P) guidelines published in(19) as shown in the flow chat (Figure 1). Primary studies that would have investigated the effectiveness of psychosocial care interventions on quality of life of adult cancer patients will be identified thorough searches in the various electronic databases that are known to produce optimal and efficient searches for systematic reviews and meta-analyses(20) that include: Ovid MEDLINE(R), PubMed, EMBASE, APA PsychINFO, Web of Science, PubMed, and Google Scholar. Studies published between 1^st^ January 2002 and 31^st^ December 2023 in any LMIC (Appendix 1), will be searched.

**Figure 1:**
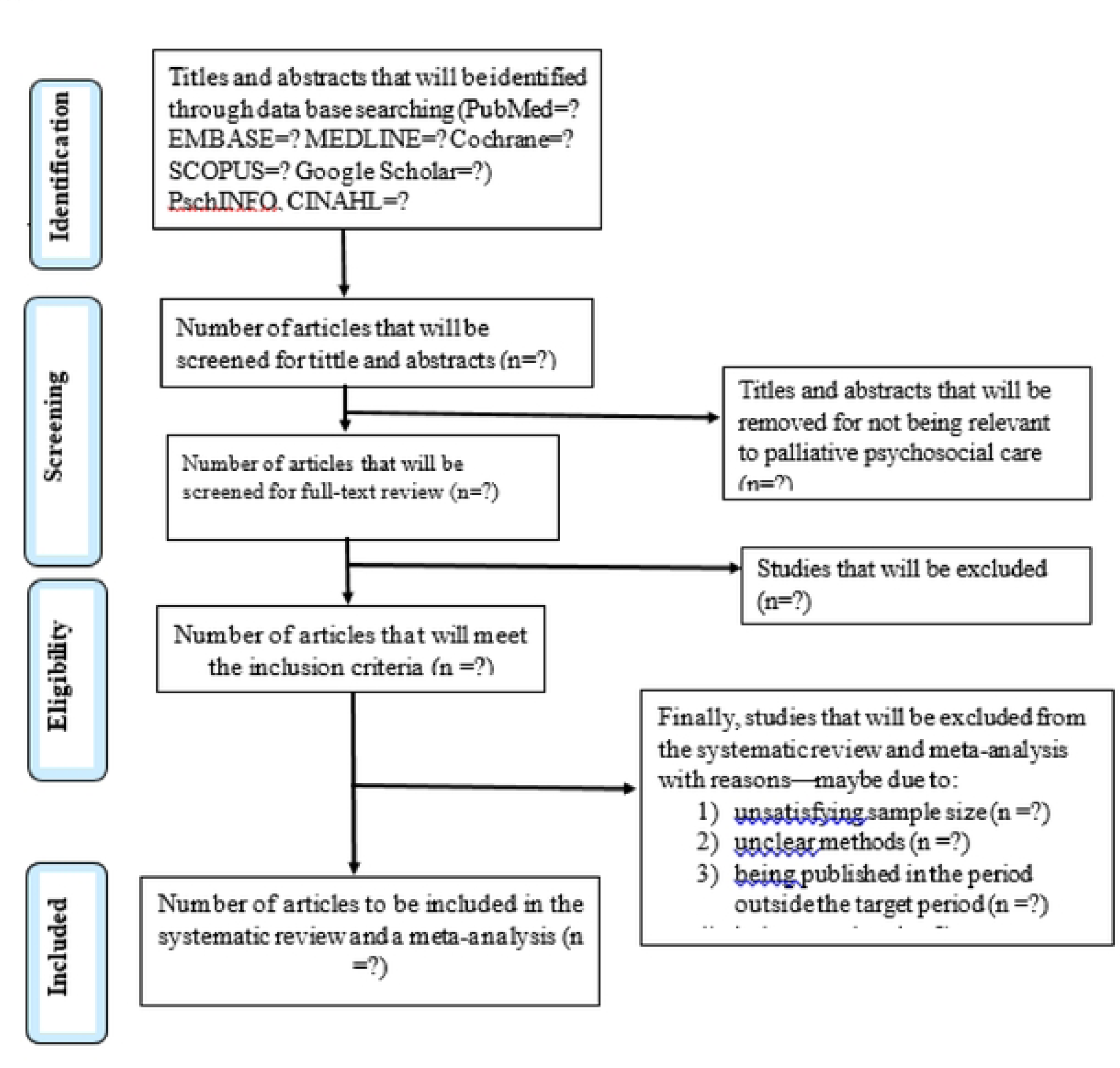
The (PIUSAIA)jlow chat summarizing ll,edata bases lobe searched, screening procedurB l1J1d /}111eligible studies/or inclusion in data synthesis

### 2.2 Search strategy

After developing the systematic review question, we developed a search strategy following the PICOST (Population, Intervention, Comparison, Outcome, Study design, Setting and Time-frame) model (19,21,22): (1) Patient or Population: adult patients with cancer in low-and middle-income countries (that is “APWC” and all related terms); (2) Intervention: Psychosocial care or Psychosocial support that focused on improving the quality of life of adult patients with cancer (that is, “PC” and all related terms); (3) Comparison: comparison between intervention/study group and control group receiving the usual cancer care; (4) Outcome: outcome from the intervention (that is, “QoL” and all related terms; (5) Study design: any randomised controlled trial study (that is “RCT”), quasi-experimental studies, cohort studies controlled before-and-after studies and other designs with comparison group.

In order to maximize the retrieval of relevant papers for our meta-analysis, we generated the key terms from the topic(19). Later, we looked for synonyms of the key terms from Medical Subject Headings (MeSH) database and relevant papers. The search strategy with the strings and their respective synonyms are included in Table 1. The search strategy will change depending on the requirements for different databases.

**Table 1:** The PICOS Model and Kev ternL for the Search Strateo•.

| PICOS items | Description | Synonyms |
| --- | --- | --- |
| Population | Adult male and female patients with any type of cancer, aged $\geq 18$ years in low- and middle-income countries. | “adult patients with cancer” OR “adult oncology patients” OR “adult with cancer” OR “women with cancer” OR “men with cancer” OR “man with cancer” OR “woman with cancer” OR “elderly with cancer” OR “middle aged with cancer” OR “aged with cancer” OR “geriatric age group with cancer” OR “geriatric oncology age group” OR “women with neoplasm” OR “woman with neoplasm” OR “elderly with neoplasm” OR “middle aged with neoplasm” OR “aged with neoplasm” OR “geriatric age group with neoplasm” OR “women with carcinoma” OR “men with carcinoma” OR “man with carcinoma” OR “woman with carcinoma” OR “elderly with carcinoma” OR “middle aged with carcinoma” OR “aged with carcinoma” OR “geriatric age group with carcinoma” |
| Intervention | Psychosocial care/psychosocial support. | “psychosocial care” OR “supportive care” OR “palliative care” OR “psychological care” OR “social care” OR “palliative and psychosocial care” OR “palliative care and psychosocial” OR “psychosocial support” OR “emotional support” OR “psychological assistance” OR “psychosocial rehabilitation” OR “mental health rehabilitation” OR “psychosocial support system” OR “social support systems” OR “psychosocial intervention” OR “psycho-oncology” OR “psychosocial oncology” OR “psychosocial palliative care” OR “psychosocial support in palliative care” OR “psychosocial wellbeing in palliative care” OR “psychosocial supportive intervention” OR “psychosocial concerns in palliative care” OR “psycho-oncological care” OR “psychosocial treatment” |
| Comparison | Comparison of the study group receiving the intervention with the control or non-intervention group. | None |
| Outcome | Improvement in physical well-being, social well-being, emotional or mental health well-being; cognitive well-being, spiritual well-being and general quality of life in the intervention group. | “quality of life” OR “quality of living” OR “well-being” OR “health-related quality of life” OR “life of quality” OR QoL |
| Study design | Randomised controlled trial (RCT); quasi-experimental studies; controlled before-and-after cohort studies, studies with comparison groups—will be eligible for inclusion. Unpublished studies, trial protocols, systematic reviews, conference abstracts will be excluded. | “randomised controlled trial” OR clinical trial OR “controlled clinical trial” OR “non-randomised controlled trial” OR “non-randomised clinical trials” OR “RCT” |
| Setting | Low-and middle-income countries (LMICs) as listed by World Bank. | Appendix 1 |
| Time-frame | 1<sup>st</sup> Jan 2002 to 31<sup>st</sup> Dec 2023 |  |

The search strategy will be amended depending on the requirements of each database.

Literature search results will be uploaded in Endnote 21 reference manager. Citation abstracts and full-text papers will be uploaded. The duplicates will be removed from the search results that will be exported to Endnote 21 software. The process of formal screening by title, abstract and full-text will follow. The Endnote reference manager will facilitate the creation of the PRISMA flow diagram once the screening process is completed.

### 2.3 Data collection

The primary reviewer (DK) will conduct the searches, do data extraction, screen the titles and abstracts yielded by the search and will appraise the quality of all papers to be included in the review. Thereafter, AAK and EAO will obtain the full-text reports and decide whether these meet the inclusion criteria. In order to reduce on the risk of bias, NN, EN, JD, SK and FEM, will check the papers included for the review. In circumstances where the consensus may not be reached, we will resolve them through a discussion with guidance of the ninth reviewer (FN).

### 2.4 Data extraction from selected papers for review and meta-analysis

The reviewers will develop an excel sheet which will be used to extract data from the included studies in the review. Thereafter, DK, AAK, EAO, NN and EN, will, in duplicate, independently use the excel sheet to extract data (Appendix 2) from the included studies. Calibration exercise will be done by the reviewers before starting the review process, to ensure consistency during data collection. For every included study, the excel sheet will extract the study author(s) and year of publication, the country where the study was conducted, study aim, study design, study participants, intervention, study period and attrition, outcome of the intervention, outcome measures and results. Other reviewers, JD, SK, FEM and FN will check for consistency and accuracy of the abstracted data by the five reviewers.

In addition, studies that will be in conformity with acceptable scientific research standards and meeting the protocol inclusion criteria, will be include for the meta-analysis. In the data extraction process, a thorough review of the title, abstract and full paper will be done by the primary reviewers while following the preferred reporting items for systematic reviews and meta-analyses (PRISMA)(19) flow chart (Figure 1). A full-text analysis of actually qualifying studies including identification of duplicated records will be conducted. Only the full-text papers will be retained for data extraction.

In a situation where a full-text is not available, the reviewers will make effort to engage the original author(s) to provide the full paper.

### 2.5 Inclusion criteria

#### Type of study participants

Adult patients aged 18 years and above, with any type of cancer, in LMICs, will be considered for this meta-analysis.

#### Type of intervention

Only psychosocial care interventions that targeted to improve the QoL of adult patients with cancer, will be included for this systematic review and meta-analysis.

#### Type of studies

This meta-analysis will consider Randomised controlled trials (RCTs); quasi-experimental studies; cohort studies controlled before-and-after studies and other designs with comparison group, comparing the effect of psychosocial care intervention. Preliminary studies to be included in the review are included in Table 3.

#### Type of outcome measures

Any objectively measured QoL (with standard tools) focusing on the physical, mental or cognitive, social, emotional, spiritual well-being and general quality of life will be considered for this systematic review and meta-analysis.

#### Setting

Studies from LMICs published in peer reviewed journals in English language between January 2002 and December 2023, will be considered for this meta-analysis.

Studies published between January 2002 and December 2023 will be searched. This is because, in 2002, the WHO redefined palliative care(8) and later, in 2014, at the Sixty-Seventh World Health Assembly (WHA67.19) resolved that all UN member countries should strengthen the integration of comprehensive palliative care (given the ever increasing cases of non-communicable diseases) in the continuum of care; that is patient and family-centered during pain and symptom management and psychosocial support provision(12,23). It is believed that, from 2002 and following the 2014 WHA resolution on strengthening palliative care, various countries (especially low, middle and high income) strengthened cancer care services in the continuum of care in order to contribute to universal health coverage for all at all levels which is the global target by the year 2030(8,12,23).

### 2.6 Exclusion criteria

Case reports, systematic reviews, meta-analysis reviews, abstracts of conferences, studies that never used case-control or cohort study design, studies with insufficient or inaccessible data in the full text, pre-prints, as well as studies that were published before 1^st^ January 2002 or after 31^st^ December 2023, will be excluded. The inclusion and exclusion criteria is summarized in Table 2.

**Table 2:** Inclusion criteria.

| Study characteristics | Inclusion criteria |
| --- | --- |
| Population | Adults patients, aged 18 years and above, with any form of cancer. |
| Intervention | Psychosocial care (individual and/or group support or chemotherapy counselling, spiritual support, psycho-education, relaxation activities, peer support, dignity therapy ...). |
| Comparison | The study group receiving the intervention with the control or non-intervention group versus the group receiving pharmacological interventions (medical, radiotherapy and surgery). |
| Outcomes | • Improvement in physical well-being.<br>• Improvement in mental or emotional well-being (reduction in stress, depression, anxiety ...).<br>• Improvement in cognitive well-being.<br>• Improvement in social well-being.<br>• Improvement in spiritual well-being.<br>• Improvement in general Quality of life. |
| Study designs | Randomised controlled trials (RCTs); quasi-experimental studies; cohort studies controlled before-and-after studies and other designs with comparison group. |
| Setting | LMICs |
| Time duration | 2002-2023 |

### 2.7 Quality assessment

The Newcastle–Ottawa scale (NOS) will be used to assess the quality of eligible publications(24). Eligible studies will be judged on three broad perspectives: the selection of study groups; the comparability of the groups; and the ascertainment of either exposure or outcome of interest for case-control studies (Appendix 3). The study with scores of 6, 7, 8 or 9 on NOS, will indicate eligibility to have satisfactorily met the quality of publication and will be considered for the meta-analysis. The four reviewers: JD, SK, FEM and FN will independently assess the extracted data from the included papers for review to assess for overall methodological quality. In case of disagreements, differences will be resolved by a consensus among all the reviewers. FN will act as the arbitrator.

Data from the research papers or their abstracts will be entered into the data extraction excel sheet on the weekly basis by the primary reviewers (DK, AAK, EAO, NN and EN). After the extraction and appraisal of the eligible studies for the systematic review and meta-analysis, every selected study will be evaluated by the other reviewers (JD, SK, FEM and FN) to validate their inclusion or exclusion in the systematic review and meta-analysis.

### 2.8 Data availability

All relevant data from this systematic review and meta-analysis will be made available upon study completion.

### 2.9 Data synthesis

### 2.10 Statistical analysis

The Odds Ratios (ORs) with the 95% Confidence Intervals (CIs) of the psychosocial intervention with improved quality of life will be evaluated among the cases and controls. Studies that will quantify the effect of the psychosocial intervention on the quality of life, the Odds of association of the intervention to the quality of life will be used. In contrast, non-quantified effect of intervention on the quality of life a descriptive account will be given.

Heterogeneity will be evaluated by calculating the heterogeneity (I^2^) statistic and a value of 50% will be used as the cut off(25). For those pooled studies with I^2^ ≥50%, random-effects model(26) will be used to pool the Odds Ratios among the cases and then controls(26). For pooled studies with I^2^<50%, the fixed effect models(27) will be used.

The significance of the pooled Odds Ratio will be determined by the Z-test, and p < 0.05 will be considered as statistically significant(28). An estimate of potential publication bias will be displayed on funnel plot in which the standard error of Odds Ratio of each study will plotted against its Odds Ratio(29). An asymmetric plot will suggest a possible publication bias and will be further assessed by the method of Egger’s linear regression test that uses the Kendall’s Rank correlation(30,31) whose significance will be determined by the t-test and a p < 0.05 will be considered representative of statistically significant publication bias. Analysis will be performed by using the statistical software MedCalc version 19.1.3.

### 2.11 Additional analyses (such as sensitivity or subgroup analyses)

Subgroup analysis will involve the different types of cancers, interventions, effects of psychosocial interventions, duration of interventions and study designs.

Sensitivity analysis will be done by removing one of the studies preferably the one with the high sample size and assess the risk of bias of that individual study. A funnel plot will be used to evaluate this. In addition, a regression analysis will be done to evaluate the effectiveness of an intervention over the given period of time.

### 2.12 Meta-biases

In order to strengthen the validity of this systematic review and meta-analysis(32), the reviewers will endeavor to address the meta-biases, that could be existing in the selected studies for review. Five reviewers (EAO, JD, SK, FEM and FN) will establish whether there were any procedural gaps during the conduct of the original study(32). Information on whether the protocol of the conducted primary study was registered and/or published before the study was started, will be explored. The funnel plots will as well be utilized to detect any possible publication bias.

For quantitative publication bias analysis, Egger’s test analysis will be used. A p>0.05 will indicate evidence of publication bias. Trim and Fill will be used to cater for the publication bias.

### 2.13 Certainty of evidence

The Grading of Recommendations, Assessment, Development and Evaluation (GRADE) guidelines(33) will be used to rate the quality of evidence from the meta-analyses. The GRADE guidelines classifies the quality of evidence in one of the four levels: high quality (further research is very unlikely to change the confidence in the estimate of intervention effect), moderate quality (further research is likely to have an important impact on the confidence in the estimate of intervention effect and may change the estimate), low quality (further research is very likely to have an important impact on the confidence in the estimate of effect and is likely to change the estimate) and very low quality (any estimate of effect is very uncertain)(33).

## 3.0 Dissemination of findings

Findings from the data synthesis will be published in a peer-reviewed journal that will be agreed upon by the reviewers. Findings from the systematic review and meta-analysis will also be presented at local and international conferences and webinars organized by African Palliative Care Association (APCA), Palliative Care Association of Uganda (PCAU) and Makerere University School of Public Health. It is believed that the findings will be utilized by palliative care or cancer care providers, policy makers and other stakeholders that to contribute towards universal health coverage (UHC) and health for all at all ages in Uganda and beyond.

## 4.0 Discussion

Available literature reiterates the significance of addressing psychosocial needs of patients with cancer(7,34). Many cancer patients tend to be desperate to try anything that would improve on their quality of life(34). This means that the cancer care providers need to be holistic in their approaches by addressing the physical, social, psychological and spiritual needs of cancer patients in order to help them have an improved quality of life. So, there is generally a global advancement in pain and symptom management among cancer patients(35) in most countries especially in chemotherapy and radiotherapy(36). Relatedly, in some countries have gone further to integrate psychosocial care interventions in care and this has yielded immense benefits for cancer patients during treatment and at or near their end of life(37). Nevertheless, available literature indicates that there is generally a slow pace in integrating psychosocial care with biomedical interventions in most low-and medium income countries which are shouldering the highest global cancer burden. Given the scanty systematic reviews and meta-analyses on the effectiveness of psychosocial care interventions in cancer care, there is a dire need for synthesising the primary studies in various contexts about the effectiveness of psychosocial care interventions on the quality of life of adult cancer patients. Consequently, the results would inform the evidence-based utilization of psychosocial care interventions for cancer patients in Uganda and beyond.

## 5.0 Conclusion

This systematic review and meta-analysis intends to determine the effectiveness of psychosocial care interventions in the improvement of the quality of life of adult patients with cancer. The results will inform future policies, research and practice in cancer care in a variety of settings.

## Author details

**DK** is a doctoral candidate at Makerere University School of Public Health in the Department of Disease Control and Environmental Health. He is a Public Health specialist and Counselling Psychologist.

**EN** is a Research manager at the African Palliative Care Association and a PhD graduate from Cicely Saunders Institute King’s College London. She specialized in Clinical Epidemiology, Biostatistics, Palliative Care, Policy and Rehabilitation during her Master’s degree training.

**NN** is a cancer researcher at the Uganda Cancer Institute working on the molecular characterization of cancers in Sub-Saharan Africa (SSA) and the development of affordable low-cost diagnostics for cancers in resource-limited settings. He completed his doctoral studies at the University of Washington where his work focused on HIV reservoir eradication and the design of PCR-based assays to quantify the latently integrated HIV. NN is currently the laboratory director and head of research and training at the Uganda Cancer Institute. He is an investigator on several studies seeking to elucidate the molecular profile of breast cancer in SSA. He is also involved in building capacity for cancer laboratory diagnostics in Uganda.

**JD** is the Chief Executive for International Children’s Palliative Care Network, visiting professor University of Belgrade, Serbia, and Edge Hill University, UK; Senior Research Fellow, Cicely Saunders Institute, Dept. of Palliative Care & Policy King’s College London; International Palliative Care Consultant.

**AAK** is Library and Information Scientist (PhD. MSc.Inf. Sc. Makerere University) in Makerere University. She heads Albert Cook Library at the College of Health Sciences, Makerere University.

**SK** is a Senior Lecturer, in the Department of Epidemiology & Biostatistics at Makerere University School of Public Health in the College of Health Sciences. He holds a PhD (Epidemiology) from the Swiss Tropical and Public Health Institute, University of Basel, a masters of science (EPI/BIO) from Case Western Reserve Univeristy and a bachelors in Statistics from Makerere University.

**FEM** is currently a Professor in the department of Epidemiology and Biostatistics-School of Public Health, College of Health Sciences, Makerere University-Kampala. He graduated with a Bachelor of Statistics (Bstat, Hons) at Makerere University Institute of Statistics and Applied Economics (ISAE) in 1993, and a MHS (2000) and PhD (2004) degrees with majors in epidemiology and population studies from the Johns Hopkins Bloomberg School of Public Health. In 2000 he was awarded The Edward J. Dehne Award in Population Dynamics, Department of Population, Family and Reproductive Health, Johns Hopkins University, Bloomberg School of Public Health.

**EAO** is a Uganda Physician, researcher, academic and health policy expert who is the immediate past president of the Uganda Medical Association, a professional industry association that champions medical doctors’ interests in Uganda. Has a PhD in Health Policy from Makerere University and McMaster University.

**FN** is a Professor of Disease Control and Prevention at Makerere University School of Public Health. He has a Bachelor of Medicine and Bachelor of Surgery degree from Makerere University; a Master’s in Public Health from Leeds University; and a PhD in Disease Control from Karolinsika Institutet. His fields of specialty and research interests include Non-Communicable Diseases (NCDs), Neglected Tropical Diseases (NTDs), HIV/STIs, TB, and Malaria.

## Author contributions

### Conceptualisation of the idea

David Kavuma.

### Methodology

David Kavuma, Alison Annet Kinengyere, Ekwaro Anthony Obuku, Eve Namisango.

### Writing original draft

David Kavuma, Ekwaro Anthony Obuku, Eve Namisango, Nixon Niyonzima

### Review and editing

David Kavuma, Eve Namisango, Julia Downing, Nixon Niyonzima, Alison Annet Kinengyere, Simon Kasasa, Fredrick Edward Makumbi, Ekwaro Anthony Obuku, Fred Nuwaha

## Data Availability

Data will be made publicly available.

## Appendices

### Appendix 1

#### Low-and Middle-income countries accordin!! to World Bank

AFGHANISTAN; ALBANIA; ALGERIA; AMERICAN SAMOA; ANGOLA; ARGENTINA; ARMENIA; AZERBAIJAN; BANGLADESH; BELARUS; BELIZE; BENIN; BHUTAN; BOLIVIA; BOSNIA AND HERZEGOVINA; BOTSWANA; BRAZIL; BULGARIA; BURKINA FASO; BURUNDI; CABO VERDE; CAMBODIA; CAMEROON; CENTRAL AFRICAN REPUBLIC; CHAO; COLOMBIA; COMOROS; CONGO, DEM. REP.; CONGO, REP.; COSTA RICA; COTE D’IVOIRE; CUBA; DJIBOUTI; DOMINICA; DOMINICAN REPUBLIC; ECUADOR; EGYPT. ARAB REP.;; EL SALVADOR; EQUATORIAL GUINEA; ERITREA; ESWATINI; ETHIOPIA; FIJI; GABON; GAMBIA. THE; GEORGIA; GHANA; GRENADA; GUATEMALA; GUINEA; GUINEA-BISSAU; GUYANA; HAITI; HONDURAS; INDIA; INDONESIA; IRAN. ISLAMIC REP.; IRAQ; JAMAICA; JORDAN; KAZAKHSTAN; KENYA; KIRIBATI; “KOREA, DEM. PEOPLE’S REP.; KOSOVO; KYRGYZ REPUBLIC; LAO PDR; LEBANON; LESOTHO; LIBERIA; LIBYA; MADAGASCAR; MALAWI; MALAYSIA; MALDIVES; MALI; MARSHALL ISLANDS; MAURITANIA; MAURITIUS; MEXICO; MICRONESIA. FED. STS.; MOLDOVA; MONGOLIA; MONTENEGRO; MOROCCO; MOZAMBIQUE; MYANMAR; NAMIBIA; NEPAL; NICARAGUA; NIGER; NIGERIA; NORTH MACEDONIA; PAKISTAN; PALAU; PAPUA NEW GUINEA; PARAGUAY; PERU; PHILIPPINES; RUSSIAN FEDERATION; RWANOA;SAMOA; SAO TOME AND PRINCIPE” OR SENEGAL OR SERBIA OR SIERRA LEONE OR SOLOMON ISLANDS OR SOMALIA; SOUTH AFRICA; SOUTH SUDAN; SRI LANKA; ST. LUCIA; ST. VINCENT AND THE GRENADINES; SUDAN SURINAME; SYRIAN ARAB REPUBLIC; TAJIKISTAN; TANZANIA; THAILAND; TIMOR-LESTE; TOGO; TONGA; TUNISIA; TURKIYE; TURKMENISTAN; TUVALU; UGANDA; UKRAINE; UZBEKISTAN; VANUATU; VIETNAM; WEST BANK AND GAZA; YEMEN, REP.; ZAMBIA; ZIMBABWE

### Appendix 2

#### Sample of Excel sheet to be used for extracting eligible studies for review

| No. | Study author(s) and year | Country | Aim(s) | Study design | Study participants | Intervention | Study period and attrition | Outcome(s) | Outcome measure(s) | Results |
| --- | --- | --- | --- | --- | --- | --- | --- | --- | --- | --- |
| 1 |  |  |  |  |  |  |  |  |  |  |
| 2 |  |  |  |  |  |  |  |  |  |  |

### Appendix 3

#### Newcastle-Ottawa Quality Assessment Scale for Case-Control studies

<u>Note:</u> A study can be awarded a maxirnum of one star for each numbered itern within the Selection and Exposure categories. A maximu1n of two stars can be given for Comparability.

##### Selection

1. Is the case definition adequate?

a. yes, with independent validation
b. yes, eg record linkage or based on self-repo11s
c. no description
2. Representativeness of the cases

a. consecutive or obviously representativeseries of cases
b. potential for selection biases or not stated
3. Selection of Controls

a. co1nn1unity controls
b. hospital controls
c. no description
4. Definition of Controls
  a. no history of disease (endpoint)
  b. no description of source

##### Comparability

1. Comparability of cases and controls on the basis of the design or analysis

a. study controls for _______ (Select the 1nost ilnportant factor.)
b. study controls for any additional factor (This criteria could be rnodified to indicate specific control for a second in1portant factor.)

##### Exposure

1. Ascertainment of exposure

a. secure record (eg surgical records)
b. structured interview where blind to case/control status
c. interview not blinded to case/control status
d. wrillen self report or 1nedical record only
e. no description
2. Same 1nethod of ascertain1nent for cases and controls

a. yes
b. no
3. Non-Response rate

a. same rate for both groups
b. non respondents described
c. rate different and no designation

#### Sample of Excel sheet to be used for scoring eligible studies for meta-analysis

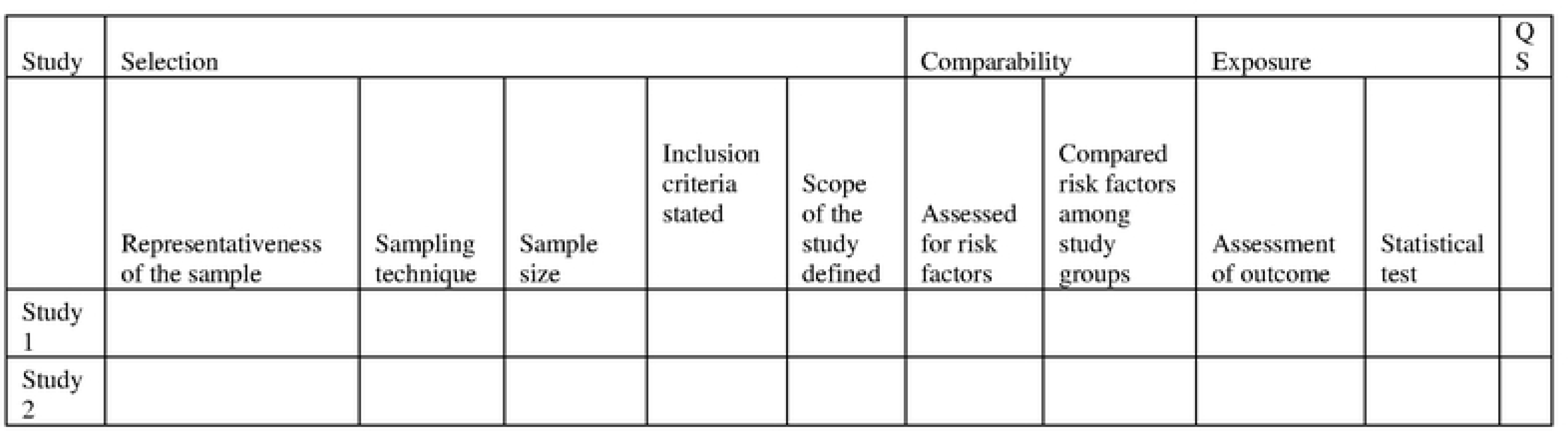

## References

1. Ferlay J, Colombet M, Soerjomataram I, Mathers C, Parkin DM, Piñeros M, et al. Estimating the global cancer incidence and mortality in 2018: GLOBOCAN sources and methods. Int J Cancer. 2019;144(8):1941–53.

2. Report by the Secretariat Executive Board WHO 140th session. Cancer prevention and control in the context of an integrated approach. 2016;22(May):1–9.

3. Grassi L, Spiegel D, Riba M. Advancing psychosocial care in cancer patients [version 1; referees: 3 approved] Referee Status: 2017;6(0).

4. Robb KA, Simon AE, Miles A, Wardle J. Public perceptions of cancer: A qualitative study of the balance of positive and negative beliefs. BMJ Open. 2014;4(7):1–6.

5. Zimmermann FF, Burrell B, Jordan J. The acceptability and potential benefits of mindfulness-based interventions in improving psychological well-being for adults with advanced cancer: A systematic review. Complement Ther Clin Pract. Elsevier Ltd; 2018;30:68–78.

6. Atlas G, Care P. Global Atlas of Palliative Care at the End of Life Global Atlas of Palliative Care. 2020.

7. Grassi L, Watson M. Psychosocial care in cancer: An overview of psychosocial programmes and national cancer plans of countries within the international federation of psycho-oncology societies. Psychooncology. 2012;21(10):1027–33.

8. Radbruch L, De Lima L, Knaul F, Wenk R, Ali Z, Bhatnaghar S, et al. Redefining Palliative Care—A New Consensus-Based Definition. J Pain Symptom Manage. Elsevier Inc; 2020;60(4):754–64.

9. African Palliative Care Association. African Palliative Care Association Standards for Providing Quality Palliative Care Across Africa. 2011;148. Available from: www.africanpalliativecare.org

10. PALLIATIVE CARE POLICY DRAFT FOR UGANDA Feb 17, 2015(2).

11. Kelley L. The World Health Organization (WHO). World Heal Organ. 2008;(July 1994):1–157.

12. Assembly SWH. Strengthening of palliative care as a component of comprehensive care throughout the life course. 2014;(May):1–5.

13. Carrasco JM, Inbadas H, Whitelaw A, Clark D. Early Impact of the 2014 World Health Assembly Resolution on Palliative Care: A Qualitative Study Using Semistructured Interviews with Key Experts. J Palliat Med. 2021;24(1):103–6.

14. Adler NE, Page AEK. Cancer care for the whole patient: Meeting psychosocial health needs. Cancer Care for the Whole Patient: Meeting Psychosocial Health Needs. 2008. 1–429 p.

15. Litwin MS. Health-Related Quality of Life.

16. Grad FP. The preamble of the constitution of the World Health Organization. Bull World Health Organ. 2002;80(12):981–2.

17. Bradley N, Lloyd-Williams M, Dowrick C. Effectiveness of palliative care interventions offering social support to people with life-limiting illness—A systematic review. Eur J Cancer Care (Engl). 2018;27(3):1–12.

18. Ã Pbj. No Title. 2009;13:6–13.

19. Page MJ, McKenzie JE, Bossuyt PM, Boutron I, Hoffmann TC, Mulrow CD, et al. Updating guidance for reporting systematic reviews: development of the PRISMA 2020 statement. J Clin Epidemiol [Internet]. Elsevier Inc.; 2021;134:103–12. Available from: 10.1016/j.jclinepi.2021.02.003

20. Bramer WM, Rethlefsen ML, Kleijnen J, Franco OH. Optimal database combinations for literature searches in systematic reviews: A prospective exploratory study. Syst Rev. Systematic Reviews; 2017;6(1):1–12.

21. Eriksen MB, Frandsen TF. The impact of patient, intervention, comparison, outcome (Pico) as a search strategy tool on literature search quality: A systematic review. J Med Libr Assoc. 2018;106(4):420–31.

22. Leonardo R. PICO: Model for Clinical Questions Evidence Based Medicine and Practice PICO: Model for Clinical Questions. Evid Based Med [Internet]. 2018;3(2):1–2. Available from: http://pubmedhh.nlm.nih.gov/nlmd/pico/piconew.php

23. International Diabetes Federation (IDF). Follow-up to the Political Declaration of the High-level Meeting of the General Assembly on the Prevention and Control of Non-communicable Diseases The. UN New York [Internet]. 2013;(27 may):66.10. Available from: http://scholar.google.com/scholar?hl=en&btnG=Search&q=intitle:Political+declaration+of+the+High-level+Meeting+of+the+General+Assembly+on+the+Prevention+and+Control+of+Non-communicable+Diseases#0

24. Stang A. Critical evaluation of the Newcastle-Ottawa scale for the assessment of the quality of nonrandomized studies in meta-analyses. Eur J Epidemiol. 2010;25(9):603–5.

25. Huedo-Medina TB, Sánchez-Meca J, Marín-Martínez F, Botella J. Assessing heterogeneity in meta-analysis: Q statistic or I 2 Index? Psychol Methods. 2006;11(2):193–206.

26. Inthout J, Ioannidis JP, Borm GF. The Hartung-Knapp-Sidik-Jonkman method for random effects meta-analysis is straightforward and considerably outperforms the standard DerSimonian-Laird method. BMC Med Res Methodol. BMC Medical Research Methodology; 2014;14(1):1–12.

27. Leonard T, Duffy JC. A Bayesian fixed effects analysis of the Mantel-Haenszel model applied to meta-analysis. Stat Med. 2002;21(16):2295–312.

28. Zaykin D V. Optimally weighted Z-test is a powerful method for combining probabilities in meta-analysis. J Evol Biol. 2011;24(8):1836–41.

29. Peters JL, Sutton AJ, Jones DR, Abrams KR, Rushton L. Contour-enhanced meta-analysis funnel plots help distinguish publication bias from other causes of asymmetry. J Clin Epidemiol. 2008;61(10):991–6.

30. Mcleod AAI. The Kendall Package. 2005;1–10.

31. Egger M, Smith GD, Schneider M, Minder C. Bias in meta-analysis detected by a simple, graphical test. Br Med J. 1997;315(7109):629–34.

32. Shamseer L, Moher D, Clarke M, Ghersi D, Liberati A, Petticrew M, et al. Preferred reporting items for systematic review and meta-analysis protocols (prisma-p) 2015: Elaboration and explanation. BMJ [Internet]. 2015;349(January):1–25. Available from: doi:10.1136/bmj.g7647

33. Guyatt GH, Oxman AD, Vist GE, Kunz R, Falck-Ytter Y, Alonso-Coello P, et al. GRADE: An emerging consensus on rating quality of evidence and strength of recommendations. Chinese J Evidence-Based Med. 2009;9(1):8–11.

34. Ullrich A, Ascherfeld L, Marx G, Bokemeyer C, Bergelt C, Oechsle K, et al. Quality of life, psychological burden, needs, and satisfaction during specialized inpatient palliative care in family caregivers of advanced cancer patients. Palliat Support Care. Springer Netherlands; 2017;16(3):1–10.

35. Evans F. Scarborough. Scarborough. 2018;68(3):182–96.

36. Albano D, Benenati M, Bruno A, Bruno F, Calandri M, Caruso D, et al. Imaging side effects and complications of chemotherapy and radiation therapy: a pictorial review from head to toe. Insights Imaging [Internet]. Springer International Publishing; 2021;12(1). Available from: 10.1186/s13244-021-01017-2

37. Rait DS. A Family-Centered Approach to the Patient with Cancer. Psychooncology. 2015;(March):561–6.

